# Assessment of diversity-based approaches used by American Universities to increase recruitment and retention of biomedical sciences faculty members: a scoping review protocol

**DOI:** 10.1101/2022.09.30.22280544

**Authors:** Britta Petersen, Sherli Koshy-Chenthittayil, Leslie A. Caromile

## Abstract

Considering the recent increased interest in diversifying the biomedical faculty workforce, a thorough assessment of the existing approaches is needed to provide guidance to universities. Hence this scoping review will be guided by the following research question, “What are the diversity-based approaches used by American Universities to increase recruitment and retention of biomedical sciences faculty members?”. The review follows the Population-Concept-Context methodology for Joanna Briggs Institution Scoping Reviews. Relevant peer-reviewed studies conducted during the last 10 years will be identified from electronic databases including Scopus, PubMed, and Embase. The search strings using keywords such as “biomedical faculty/researcher,” “hiring/retention,” and “diversity/minority/underrepresented” will be conducted using Boolean logic. Two independent reviewers will conduct all title and abstract screening, followed by a full article screening and data extraction. Due to the possible heterogeneity of the studies, we hope to use either a narrative analysis and/or descriptive figures/tables to depict the results. The review will present various diversity programs that have been implemented and evaluated in American Universities to increase the recruitment and retention of biomedical sciences faculty.

## Introduction

The need for more innovative approaches to diversify biomedical sciences faculty is evident in the changing demographics of the student body at American universities. While the national percentage of historically underrepresented college students (undergraduate and graduate students combined) has risen to nearly 50%, the percentage of historically underrepresented faculty remains below 30% ^1-5^, and the percentage of historically underrepresented tenure-track faculty remains even lower at 22% (NCES, 2018). These numbers represent the racial and ethnic categories and do not include a breakdown of other underrepresented groups, i.e., LGBTQ, veterans, and gender minorities. The benefits of a more diverse faculty extend to all students^6^. For students from traditionally underrepresented groups, engaging faculty role models from similar backgrounds sends a powerful message of support and belonging ^5^, and students from majority backgrounds gain by experiencing broader pedagogical perspectives^7^ and countering stereotypes to reduce bias^8^.

Research shows that diverse teams working together and capitalizing on innovative ideas and distinct perspectives outperform homogenous teams. Scientists from diverse backgrounds and life experiences representing the national population bring different perspectives, creativity, and individual enterprise to address complex scientific problems^9^. In addition, culturally diverse research teams are more likely to avoid biased outcomes in preclinical research and clinical trials, which traditionally focus primarily on white populations of European descent to the exclusion of other races and ethnicities^10^. However, for this to happen, one must feel comfortable working in an environment where they feel seen, heard, fairly supported, and welcomed - not separate. For these reasons and many others, successful initiatives to increase the number of faculty from underrepresented backgrounds are critical.

The academy has been discussing strategies to improve equity in the biomedical sciences for decades, but progress has been incremental and slow. There is no “one-size fits all” solution. Different academic institutions and different units within those institutions often need different approaches^11^. Unfortunately, many successful approaches are not published in peer-reviewed academic journals. This makes it challenging to create successful, diversity-centered policies. Here we present a protocol of a scoping review of the assessment of diversity-based approaches used by American Universities to increase the recruitment and retention of biomedical sciences faculty members. This scoping review aims to guide universities in bridging the gap between intent and implementation of these important initiatives.

## Methods

The stages of the review include^12^: (1) identifying the research questions; (2) identifying the relevant studies; (3) study selection; (4) charting the data; (5) collating, summarizing, and reporting the results; and (6) consultation.

### 1. Identifying the research question

Based on the Population, Concept, Context ^12^(PCC) framework put forward by JBI, the research question for this scoping review is “What are the diversity-based approaches used by American Universities to increase recruitment and retention of biomedical sciences faculty members?”. The PCC with regards to our review is shown in Table 1.

**Table 1:**
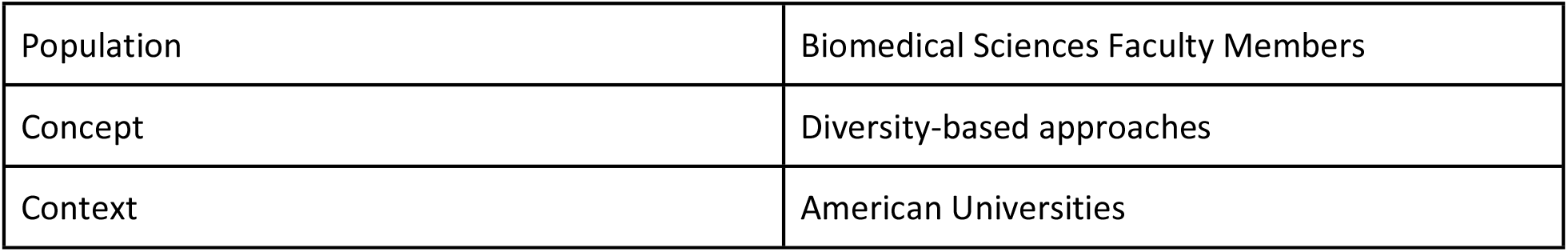
Population-Concept-Context Methodology

### (2) identifying the relevant studies

The main keywords were developed from the title, and further subcategories were derived based on association or synonyms for the topic of interest, for example, Table 2 provides the results of a pilot search in the Pubmed database.

**Table 2:** Results of a pilot search

| Keywords used | Date of search | Database | Number of results |
| --- | --- | --- | --- |
| ("Faculty"[Mesh] OR faculty[tw] OR researcher[tw]) AND biomedical[tw] AND (hiring[tw] OR retention[tw]) OR mentoring[tw]) AND (minority[tw] OR diversity[tw] OR underrepresented[tw]) | June 1, 2022 | Pubmed | 100 |

### (3) Selection of studies

Two independent reviewers will screen the records from the databases utilized using the title and abstract. The reviewers will then assess the full-text articles and will decide based on the inclusion/exclusion criteria given below.

Any disagreements will be discussed between the two reviewers until a consensus is reached, or a third reviewer will be a tiebreaker. Reasons for exclusion will be noted, and the process of study selection will be documented in a flow chart (Fig 1), according to Preferred Reporting Items for Systematic reviews and Meta-Analyses extension for Scoping Reviews (PRISMA-ScR)^13^.

**Fig 1:**
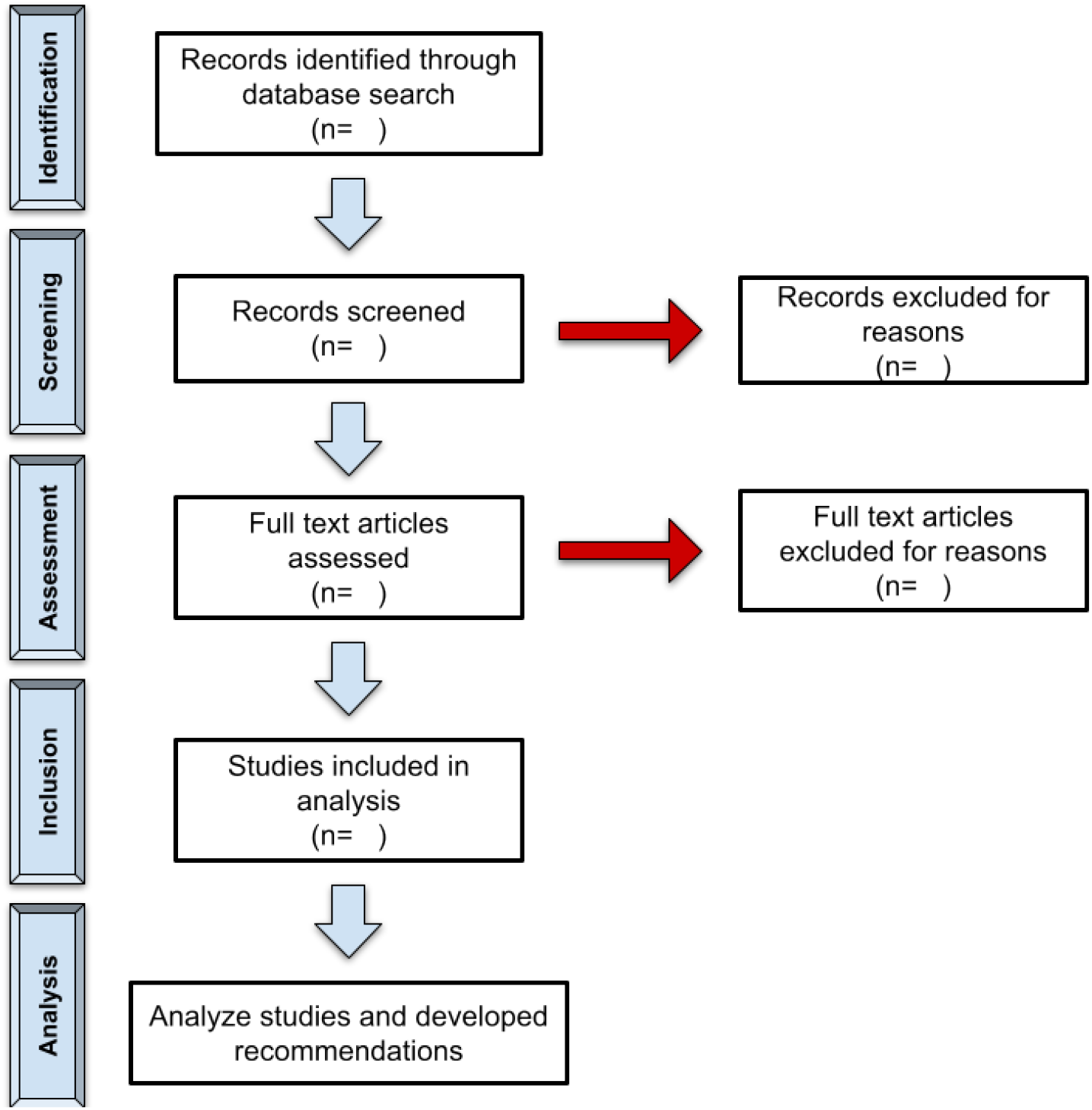
Selection of studies according to PRISMA-ScR protocol.

Eligibility criteria for this study will be as below:

#### Inclusion Criteria for This Study

- Articles published in the last 10 years but before June 1, 2022, when the search was performed
- An original intervention or observation with outcomes or evaluation data on retention, recruitment, or prevalence of historically underrepresented faculty
- Published in PUBMED, SCOPUS, EMBASE indexed journals
- Articles that focused on historically underrepresented faculty recruitment, retention, and mentoring
- Qualitative studies with pre- and post-intervention data.
- Written in English

#### Exclusion Criteria for This Study

- Papers that were narrative reviews, expert opinions, editorials, or letters to the editor
- Papers that were written more than 10 years prior to the search date
- Papers that did not include any data with their description
- Books or book chapters
- Websites that provide unpublished, non-peer-reviewed internal statistics

### (4) charting the data

We will extract the following information from the selected studies: authors, title, year of publication, the database used, the diversity-based approach used to increase recruitment and retention of biomedical faculty, number of participants in the study, and the results of the evaluation of the approach. We will contact the authors of the included papers if pertinent information is missing or unclear. To ensure consistency, the data will be extracted by one reviewer and validated by another. The data charting will be updated iteratively based on the studies found.

### (5) collating, summarizing, and reporting the results

Since this is a scoping review, the results of the data extraction will provide an overview of the different approaches adopted by American universities to increase the recruitment and retention of diverse biomedical faculty. Due to the possible heterogeneity of the studies, we hope to use either a narrative analysis and/or descriptive figures/tables to depict the results.

### (6) consultation

The results will be consulted with the relevant stakeholders (faculty members, university diversity, equity and inclusion councils, policy-decision makers, and deans).

## Discussion

College faculty have become more racially and ethnically diverse but remain far less so than students^14^. When looking at the biomedical sciences, the following racial and ethnic groups have been shown by both the National Institutes of Health and the National Science Foundation to be underrepresented: Blacks or African Americans, Hispanics or Latinos, American Indians or Alaska Natives, Native Hawaiians, and other Pacific Islanders. Additionally, women have been shown to be underrepresented in doctorate-granting research institutions at senior faculty levels in most biomedical-relevant disciplines^15^.

Although many diversity-based initiatives that aim to increase the aforementioned groups have been locally successful, these approaches are not published in peer-reviewed academic journals but instead presented in gray papers. While gray papers can be helpful in some situations, they are not peer-reviewed, frequently do not include methodological descriptions that facilitate evaluating quality, nor are included in well-curated databases of academic disciplines. Thus, making it difficult to search for and retrieve relevant information^16-18^. This scoping review will provide information on the assessment of peer-reviewed, diversity-based approaches used by American Universities to increase the recruitment and retention of underrepresented biomedical sciences faculty members.

The strengths of the scoping review will include original, peer-reviewed interventions or observations with outcomes or evaluation data on retention, recruitment, or prevalence of historically underrepresented faculty; information focusing on historically underrepresented faculty recruitment, retention, and mentoring; and qualitative studies with pre- and post-intervention data. However, there are several potential limitations of the scoping review, including that faculty underrepresentation can vary from setting to setting and, therefore, different approaches may be needed^11^. Despite the limitation, to our knowledge, the results will assist universities in bridging the gap between intent and implementation of these important initiatives.

## Data Availability

No datasets were generated or analyzed during the current study. All relevant data from this study will be made available upon study completion.

## Notes

### Competing Interest Statement

The authors have declared no competing interest.

### Clinical Trial

NA

### Funding Statement

The author(s) received no specific funding for this work

